# Reclassification of Genetic Variants in Patients with Hypertrophic Cardiomyopathy from the Sarcomeric Human Cardiomyopathy Registry (SHaRe)

**DOI:** 10.64898/2026.08.05.26359735

**Authors:** Sophie Hespe, George Powell, Laura Catto, Natalie Stewart, Amy Baker, Neesha Krishnan, Lucas A Mitchell, Natasha Henden, Ebony Richardson, Alexandra Butters, Pantazis Theotokis, Rachel Buchan, Kathryn A. McGurk, Brian Claggett, Dominic Abrams, Euan Ashley, Victoria N Parikh, Sharlene M Day, Adam S Helms, Rachel Lampert, Kim Y Lin, Joseph W. Rossano, Peter Paul Zwetsloot, Michelle Michels, Erin M Miller, Francesca Girolami, Iacopo Olivotto, Anjali Owens, Alexandre C Pereira, Thomas D Ryan, Sara Saberi, Mark W. Russell, John C Stendahl, Belinda Gray, Alessia Argiro, Niccolo Maurizi, Lia Crotti, Christoffer R Vissing, Neal K Lakdawala, Carolyn Y Ho, James S. Ware, Jodie Ingles

**Author notes:** **ADDRESS FOR CORRESPONDENCE (co-corresponding):** Associate Professor Jodie Ingles Garvan Institute of Medical Research, 384 Victoria Street Darlinghurst, Sydney, New South Wales 2010 Australia, Sophie Hespe, Garvan Institute of Medical Research, 384 Victoria Street Darlinghurst, Sydney, New South Wales 2010 Australia.

## Abstract

**Background:** Genetic testing is a Class I recommendation for patients with hypertrophic cardiomyopathy (HCM). Variant classification relies on evidence from publicly available case data, evolving classification rules, and gene-disease associations. Thus, as knowledge increases, genetic variant classifications change over time. We evaluated the occurrence and reasons for variant reclassification from a large multi-center international HCM registry (Sarcomeric Human Cardiomyopathy Registry; SHaRe), with the goal to minimize uncertainty for patients and clinicians.

**Methods:** Participants receive clinical care at specialized HCM centres. Baseline classifications were derived from the clinical genetic test report (original or updated) or prior further adjudication by SHaRe geneticists. All variants were then computationally reannotated and reevaluated during 2024-2025. Variants underwent expedited curation if no new evidence was present. The remainder underwent full manual curation using accepted criteria and classified as pathogenic/likely pathogenic (P/LP), variant of uncertain significance (VUS) and benign/likely benign (B/LB). VUS were sub-classified to high, mid or low.

**Results:** Of 12,187 HCM patients, 8,054 (66%) had genetic testing between 1990-2024, and 4,923 (61%) had a variant identified in one of 29 ClinGen-validated HCM genes (1606 unique variants). Expedited curation was performed for 704 (44%) variants and 902 (56%) underwent manual curation. There were 1279 (79%) variants that retained their classification: 148 B/LB, 663 VUS, and 468 P/LP. While 276 (17%) variants (n=557 patients) were reclassified, including 73 upgrades: 61 from VUS to P/LP (199 patients), and 12 from B/LB to VUS. There were 203 downgrades: 108 from P/LP to VUS (196 patients), and 95 from P/LP or VUS to B/LB. VUS were additionally subclassified: 90 VUS-High, 129 VUS-Mid, 115 VUS-Low. Sub-classification of VUS resulted in less uncertainty, with 369 (40.6%) variants reclassified as VUS-Low or B/LB, indicating a very strong probability of not being HCM associated.

**Conclusions:** Clinically meaningful reclassification occurred in 10% of variants identified in HCM probands. Most VUS were unlikely to be causal, and sub-classification has potential to reduce their burden on clinicians and families. Contemporary approaches to classification can minimize uncertainty of genetic results and highlight the need for periodic reevaluation.

## INTRODUCTION

Hypertrophic cardiomyopathy (HCM) affects ∼1 in 500 people^1^ and is characterized by left ventricular (LV) hypertrophy in the absence of abnormal loading conditions.^2,3^ HCM is genetically heterogeneous, with evidence for gene-disease association now spanning 29 genes from 12 ontologies.^4^ Genetic testing is a class 1 recommendation in all recent clinical management guidelines,^2,3,5,6^ as it is important for clarifying risk for family members, enabling reproductive genetic testing options,^7^ clarifying diagnoses and increasingly, for guiding clinical management.^8,9^

Variant classification involves systematically evaluating the available evidence to determine the likelihood of a genetic variant’s role in causing monogenic disease. Evidence is derived from population databases, clinical cases, family segregation, computational *in silico* scores, functional evidence, and other literature.^10^ Criteria specified by the American College of Medical Genetics and Genomics and Association for Molecular Pathology (ACMG/AMP), including recent gene-specific modifications, are commonly used.^11^ Variants are classified as benign (B), likely benign (LB), variant of uncertain significance (VUS), likely pathogenic (LP) or pathogenic (P). P and LP variants have 90% certainty of causation of disease^12^ and are clinically actionable meaning they are reported to patients, can guide clinical management, and inform cascade testing of relatives. Approximately 80% are VUS, often due to their rarity and limited functional or segregation data.^12^ Assignment of VUS spans a continuum of evidence for pathogenicity, from almost LB to almost LP. Recently, sub-classification of VUS has proven useful in capturing this nuance and better conveying the perceived importance of the variant.^8,13–15^

Previously reported variant reclassification rates for cardiac genes range from 14%^16^ to 59%^17^, with differences due to timeframes and assessment conditions. The need for periodic variant reclassification in HCM was highlighted over a decade ago,^18^ however is logistically challenging in practice. Indeed, the only study published since, from a single site cohort of 1313 HCM patients recruited between 2000-2021 showed that 22% of variants were reclassified.^19^ Taken together, evolving knowledge, gene-disease relationships and classification criteria necessitate periodic reclassification of genetic variants. While genetic testing is currently considered a ‘one-and-done’ test with periodic reclassification, the rapid pace of evolving knowledge means this approach likely needs to be reconsidered. Ultimately, genetic testing and its results are not static. Here, we report reclassification of genetic variants in a large comprehensive international HCM cohort, the Sarcomeric Human Cardiomyopathy Registry (SHaRe), and evaluate the factors driving these changes. We additionally propose a framework for reclassification of genetic variants in HCM, with the goal to minimize genetic uncertainty.

## METHODS

### The Sarcomeric Human Cardiomyopathy Registry (ShaRe)

SHaRe is an international network of specialized HCM centers, including 17 sites across 12 countries. Each center collects and maintains comprehensive longitudinal genetic, phenotypic and outcome data on HCM patients and their families. SHaRe includes ∼12,000 HCM patients and the processes, method of data collection and data curation have previously been described.^20^ Patients were included if they were probands or family members diagnosed with HCM, not including LV hypertrophy (LVH) as part of syndromic disease, and had undergone genetic testing. HCM was diagnosed by each SHaRe site and is defined by LVH, with a maximal LV thickness exceeding 15mm or an equivocal LV wall thickness z score in pediatric patients, or over 13mm in family members in the absence of abnormal loading conditions. Participation in SHaRe was independently approved by the institutional ethics board at each center, including de-identified data.

### Genetic testing

Genetic testing was performed between 1990 and 2024 using different platforms and HCM panels, reflecting the historic evolution of HCM genetic testing over time. While data from multigene panels have not been consistently captured for every patient, sequencing of 8 sarcomere genes (*MYH7*, *MYBPC3*, *TNNI3*, *TNNT2*, *TPM1*, *ACTC1*, *MYL2*, and *MYL3*) was the minimum requirement for genetically tested patients in SHaRe. Baseline variant classifications were assigned by the clinical laboratory who did the testing. Over time, some variants were reclassified by the clinical laboratory and updated in SHaRe when the patient returned to the site for cardiac review. Other variants underwent periodic adjudication by the ShaRe variant curation team, often to deal with discordant classifications. While most variants are now classified using the 5-tier classification system,^11^ many historic variants were reclassified over time for data consistency.

### Variant annotation and data cleaning

Variants were extracted from SHaRe and HGVSc notations were converted to GRCh38 coordinates, yielding unique variant IDs. Each unique variant was annotated using Ensembl Variant Effect Predictor (VEP; version 114) with ENSEMBL transcripts, corresponding HGVSc and HGVSp, VEP consequence, VEP impact, allele frequencies in population databases, in silico scores, ClinVar assertions (including, number, classification, and phenotype), SHaRe allele count, number of SHaRe carriers, number of SHaRe sites, SHaRe baseline classification. Genetic transcripts were not historically collected, and the Ensembl canonical transcript was inferred for 74% of variants. Interrogation of the data by matching the converted GRCh38 HGVSc back to SHaRe HGVSp was performed to ensure nucleotide and protein positions were as expected.

### Standardized variant reclassification

Classifications grouped for simplification, resulting in three categories: B/LB, VUS, and P/LP. Each variant was systematically reevaluated and filtered into 1 of 3 groups based on the method of curation applied: expedited variant re-curation, manual curation and curation of variants in genes of uncertain significance.

#### 1. Expedited variant re-curations

Variants underwent expedited re-curation if no new evidence sufficient to change classification was observed following reannotation. This included those with non-conflicting classifications; i.e., submitted by ≥3 SHaRe sites and ≥4 non-conflicting 3-star ClinVar assertions that are concordant with the SHaRe classification. As well as non-conflicting single submitter classifications submitted by a single SHaRe site but with ≥4 non-conflicting 3-star ClinVar assertions that were concordant with the SHaRe classification.

#### 2. Manual variant re-curations

Variants underwent manual curation if there was new evidence to change classification observed from the reannotation data, or when the variant had an absent or conflicting classification. Full manual variant curations were performed by seven independent cardiac genetic counselors and variant curators with expertise in HCM and variant classification (SH, NH, NK, AB, LC, NS, LM). Any variants that exceeded a gnomAD v4.1^21^ population allele frequency rarity threshold of ≥ 0.00004, as per accepted HCM disease prevalence (PM2_supporting, BA1, BS1, and BS2)^1,22^ were marked for manual curation. Variants in genes (*MYH7*, *MYBPC3*, *TNNI3*, *TNNT2*, *TPM1*, *ACTC1*, *MYL2*, and *MYL3*) with Clinical Genome Resource Cardiomyopathy Expert Panel (ClinGen CMP-EP) gene specific adaptations of the ACMG/AMP variant interpretation guidelines were assessed according to the gene specific criteria.^23,24^ PS4 was applied only when PM2_supporting was met using proband count thresholds; PS4_supporting n=2, PS4_moderate n=8, PS4_strong n=16.

Variants that were classified as VUS were further sub-classified into VUS-High, VUS-Mid, and VUS-Low using the Bayesian points adaptation of the ACMG criteria,^12^ in which a point scale is applied dependent on the strength of evidence; supporting +1 and −1, moderate +2 and −2, strong +4 and −4, very strong +8 and −8, for pathogenic and benign criteria relatively. The total points for each variant correspond to a final classification where: ≥ 6 points = P/LP,4 to 5 points = VUS-High, 2 to 3 points = VUS to Mid, −1 to 1 points = VUS-Low, and ≤ −2 points = B/LB.

#### 3. Curation of variants in genes of uncertain significance

As per the ACMG/AMP criteria,^11^ variants reported in genes other than the 29 genes with an established clinically significant genotype-phenotype association with HCM^4^ were classified as VUS for HCM. Additionally, for genes with established molecular mechanisms for HCM, variants that result in an alternate mechanism were classified as VUS for HCM, e.g., non-loss-of-function variants in *ALPK3*.

## RESULTS

### Genetic variants in SHaRe

Of 12,187 patients with HCM, 8054 (66%) underwent genetic testing between 1990 and 2024. Of patients with genetic testing, 4923 patients (61%; mean age at diagnosis 39.7 ± 18.3 years, 44.8% female) had a variant identified in an HCM gene, yielding 1606 unique variants (Table 1) included in this study. Most variants were in sarcomere genes (78%), with fewer in other genes associated with monogenic HCM (8%) or syndromic conditions with isolated LVH/HCM genocopies (14%). The original variant classifications included: 584 (36%) P/LP, 810 (50%) VUS, 161 (10%) B/LB variants, and 51 (3%) with absent or conflicting classifications.

**Table 1.**
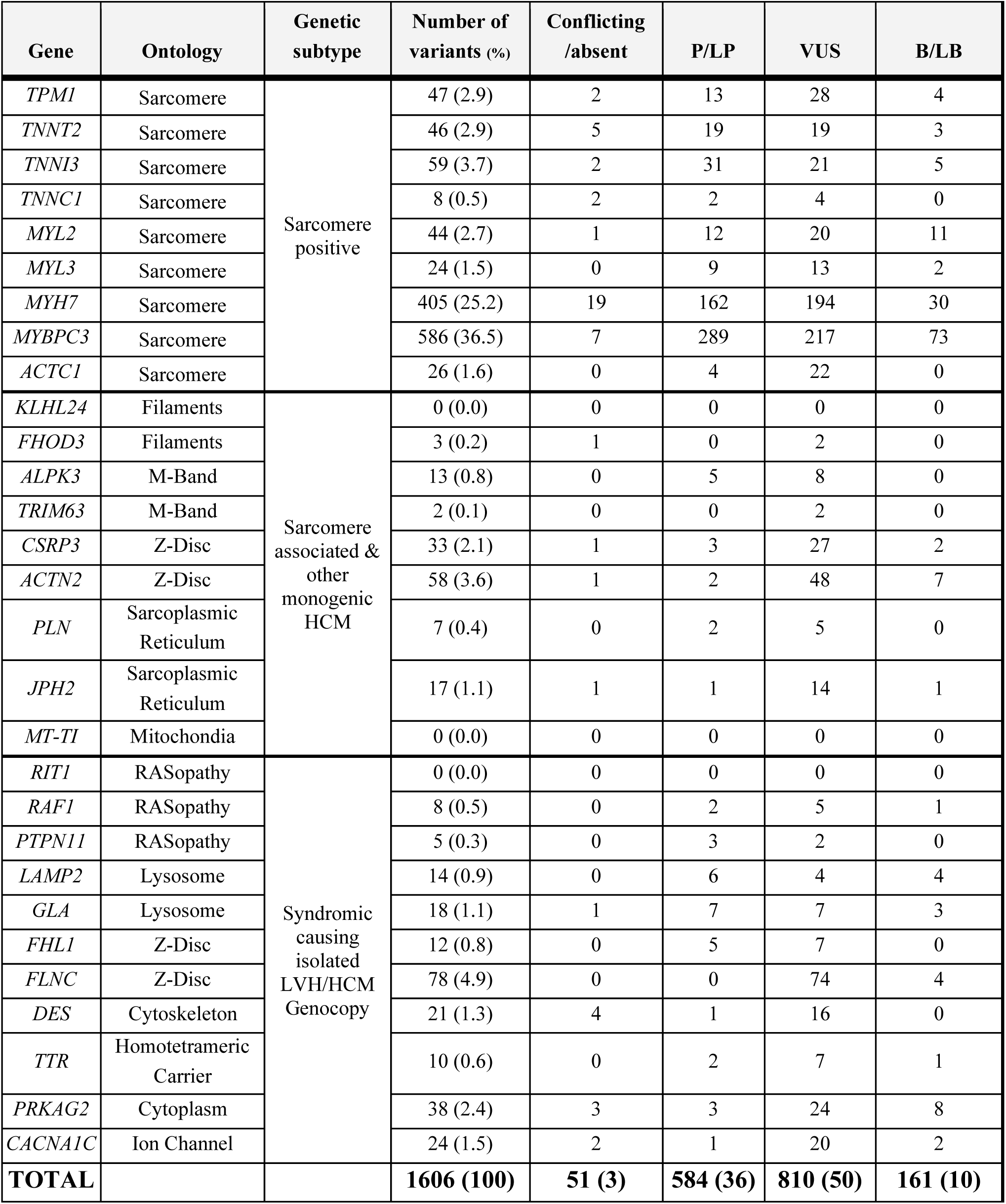
Baseline characteristics of genetic variants in HCM genes.

| Gene | Ontology | Genetic subtype | Number of variants (%) | Conflicting /absent | P/LP | VUS | B/LB |
| --- | --- | --- | --- | --- | --- | --- | --- |
| *TPM1* | Sarcomere | Sarcomere positive | 47 (2.9) | 2 | 13 | 28 | 4 |
| *TNNT2* | Sarcomere |  | 46 (2.9) | 5 | 19 | 19 | 3 |
| *TNNI3* | Sarcomere |  | 59 (3.7) | 2 | 31 | 21 | 5 |
| *TNNC1* | Sarcomere |  | 8 (0.5) | 2 | 2 | 4 | 0 |
| *MYL2* | Sarcomere |  | 44 (2.7) | 1 | 12 | 20 | 11 |
| *MYL3* | Sarcomere |  | 24 (1.5) | 0 | 9 | 13 | 2 |
| *MYH7* | Sarcomere |  | 405 (25.2) | 19 | 162 | 194 | 30 |
| *MYBPC3* | Sarcomere |  | 586 (36.5) | 7 | 289 | 217 | 73 |
| *ACTC1* | Sarcomere |  | 26 (1.6) | 0 | 4 | 22 | 0 |
| *KLHL24* | Filaments | Sarcomere associated & other monogenic HCM | 0 (0.0) | 0 | 0 | 0 | 0 |
| *FHOD3* | Filaments |  | 3 (0.2) | 1 | 0 | 2 | 0 |
| *ALPK3* | M-Band |  | 13 (0.8) | 0 | 5 | 8 | 0 |
| *TRIM63* | M-Band |  | 2 (0.1) | 0 | 0 | 2 | 0 |
| *CSRP3* | Z-Disc |  | 33 (2.1) | 1 | 3 | 27 | 2 |
| *ACTN2* | Z-Disc |  | 58 (3.6) | 1 | 2 | 48 | 7 |
| *PLN* | Sarcoplasmic Reticulum |  | 7 (0.4) | 0 | 2 | 5 | 0 |
| *JPH2* | Sarcoplasmic Reticulum |  | 17 (1.1) | 1 | 1 | 14 | 1 |
| *MT-TI* | Mitochondria |  | 0 (0.0) | 0 | 0 | 0 | 0 |
| *RIT1* | RASopathy | Syndromic causing isolated LVH/HCM Genocopy | 0 (0.0) | 0 | 0 | 0 | 0 |
| *RAF1* | RASopathy |  | 8 (0.5) | 0 | 2 | 5 | 1 |
| *PTPN11* | RASopathy |  | 5 (0.3) | 0 | 3 | 2 | 0 |
| *LAMP2* | Lysosome |  | 14 (0.9) | 0 | 6 | 4 | 4 |
| *GLA* | Lysosome |  | 18 (1.1) | 1 | 7 | 7 | 3 |
| *FHL1* | Z-Disc |  | 12 (0.8) | 0 | 5 | 7 | 0 |
| *FLNC* | Z-Disc |  | 78 (4.9) | 0 | 0 | 74 | 4 |
| *DES* | Cytoskeleton |  | 21 (1.3) | 4 | 1 | 16 | 0 |
| *TTR* | Homotetrameric Carrier |  | 10 (0.6) | 0 | 2 | 7 | 1 |
| *PRKAG2* | Cytoplasm |  | 38 (2.4) | 3 | 3 | 24 | 8 |
| *CACNA1C* | Ion Channel |  | 24 (1.5) | 2 | 1 | 20 | 2 |
| **TOTAL** |  |  | **1606 (100)** | **51 (3)** | **584 (36)** | **810 (50)** | **161 (10)** |

**Table 2.**
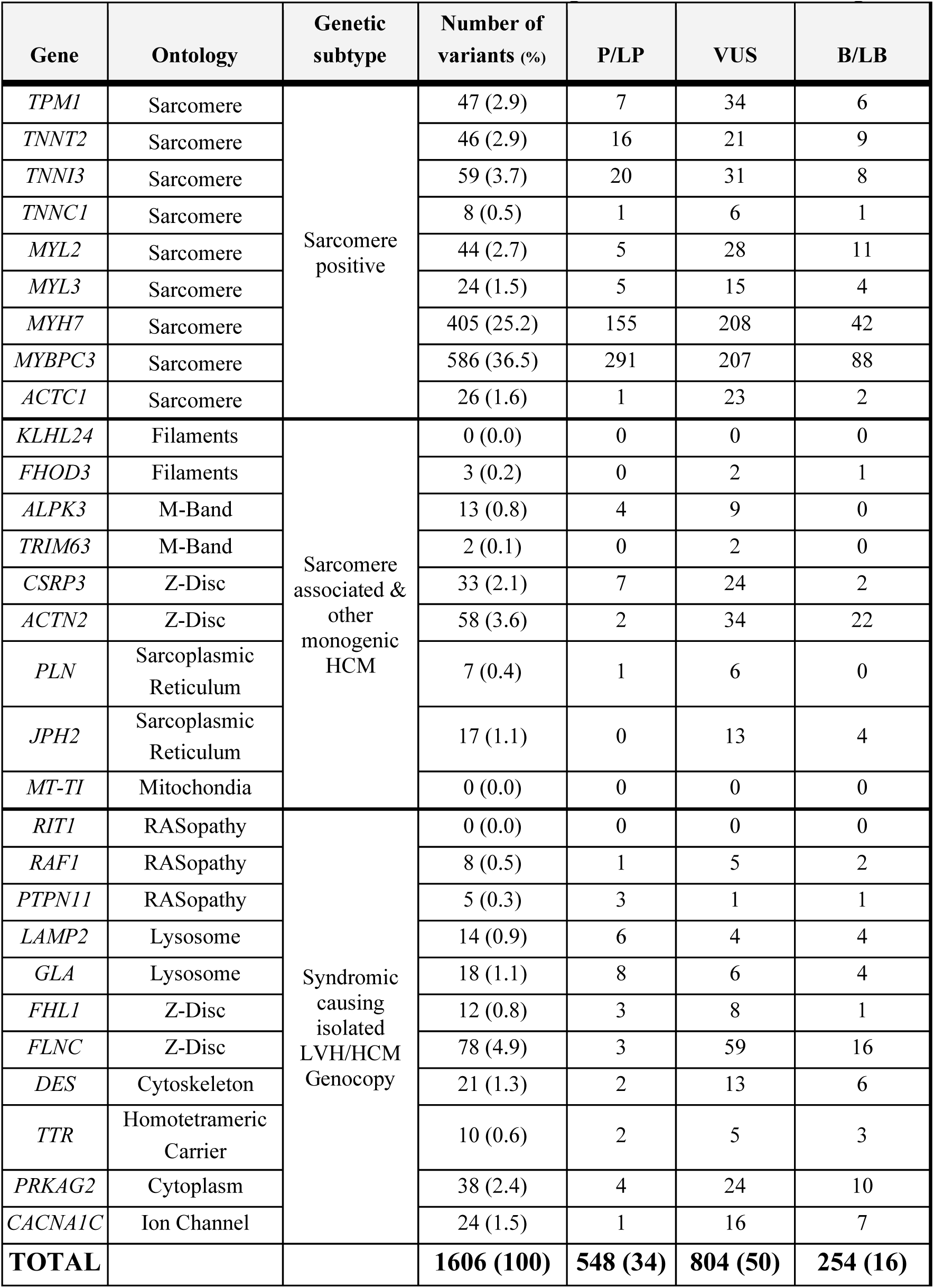
Characteristics of final reclassification of genetic variants in HCM genes.

### Variant reclassification

There were 704 (44%) variants that underwent expedited curation and 902 (56%) were manually curated (Table S1). Following reclassification, 1275 (79%) variants retained their classifications, including 146 B/LB, 660 VUS, and 468 P/LP (Figure 1A). There were 276 (17%) variants that were reclassified. Among these, 73 (26%) were upgraded and 203 (74%) downgraded from their original SHaRe classification (Figure 1B). Clinically significant reclassifications were observed for 177 (11%) variants, either upgraded to, or downgraded from P/LP, impacting 395 (5%) individuals enrolled in SHaRe who had undergone genetic testing (Figure 1B).

**Figure 1.**
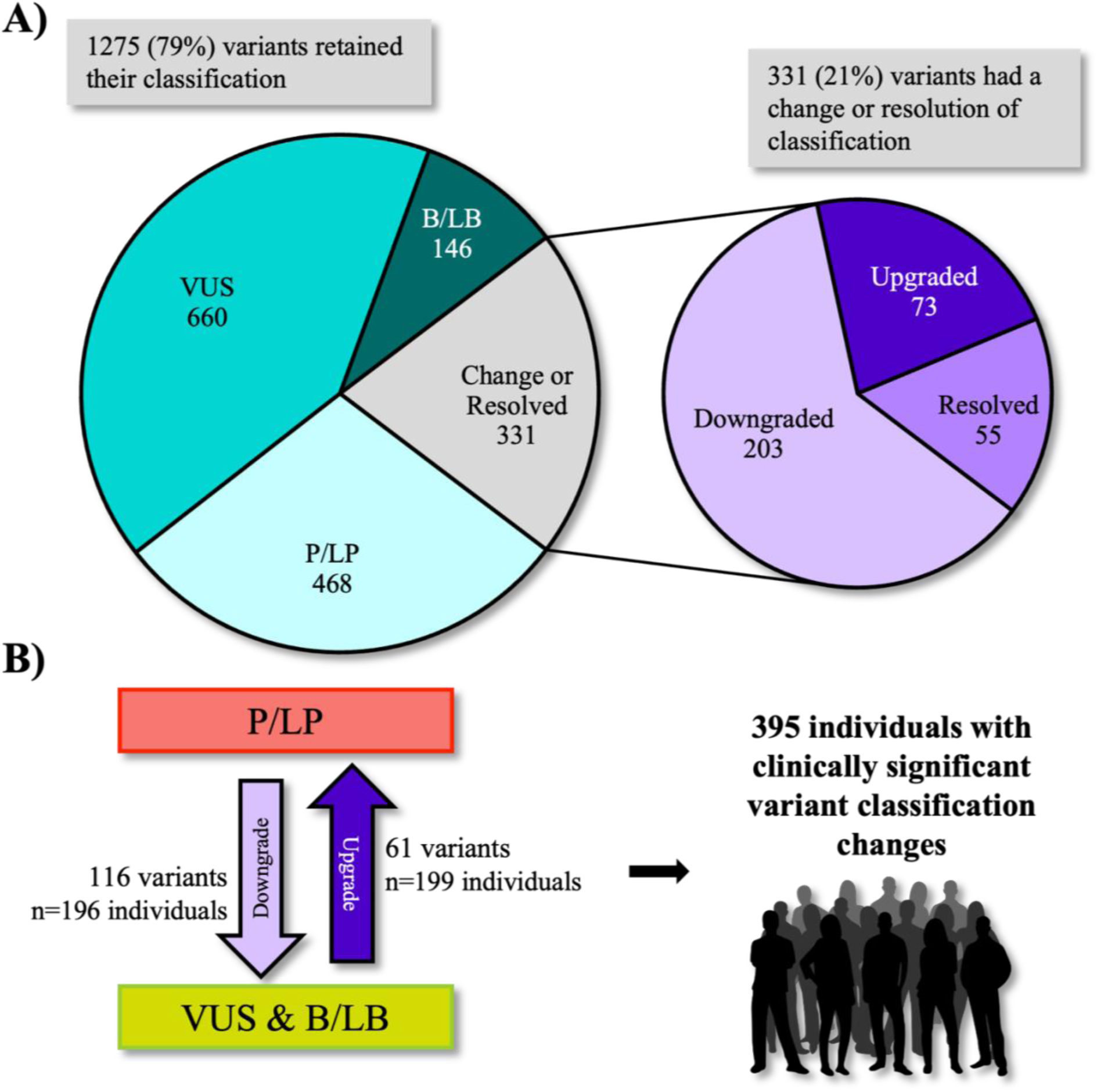
Re-curated classifications of variants compared to original classification. **A)** Proportion of variants retaining P/LP, VUS, and B/LB with total variants with a changed or resolved classification (Left); Proportions of type of change or resolved (Right). **B)** Summary of variants with clinically significant changes of classification and individuals affected. *Abbreviations: P/LP, Pathogenic/Likely Pathogenic; VUS, Variant of Uncertain Significance; B/LB, Benign/Likely Benign*.

Of the 902 manually curated variants, 334 were considered VUS. Further sub-classification of these variants included 90 (27%) VUS-High, 129 (39%) VUS-Mid, and 115 (34%) VUS-Low (Figure 2). There were 55 variants that were missing a classification or had discordant classifications in SHaRe, due to ongoing recruitment and data cleaning practices these were yet to be resolved. Of these, 31 variants had discordance resolved (Table S2) and 24 had their missing classification updated.

**Figure 2.**
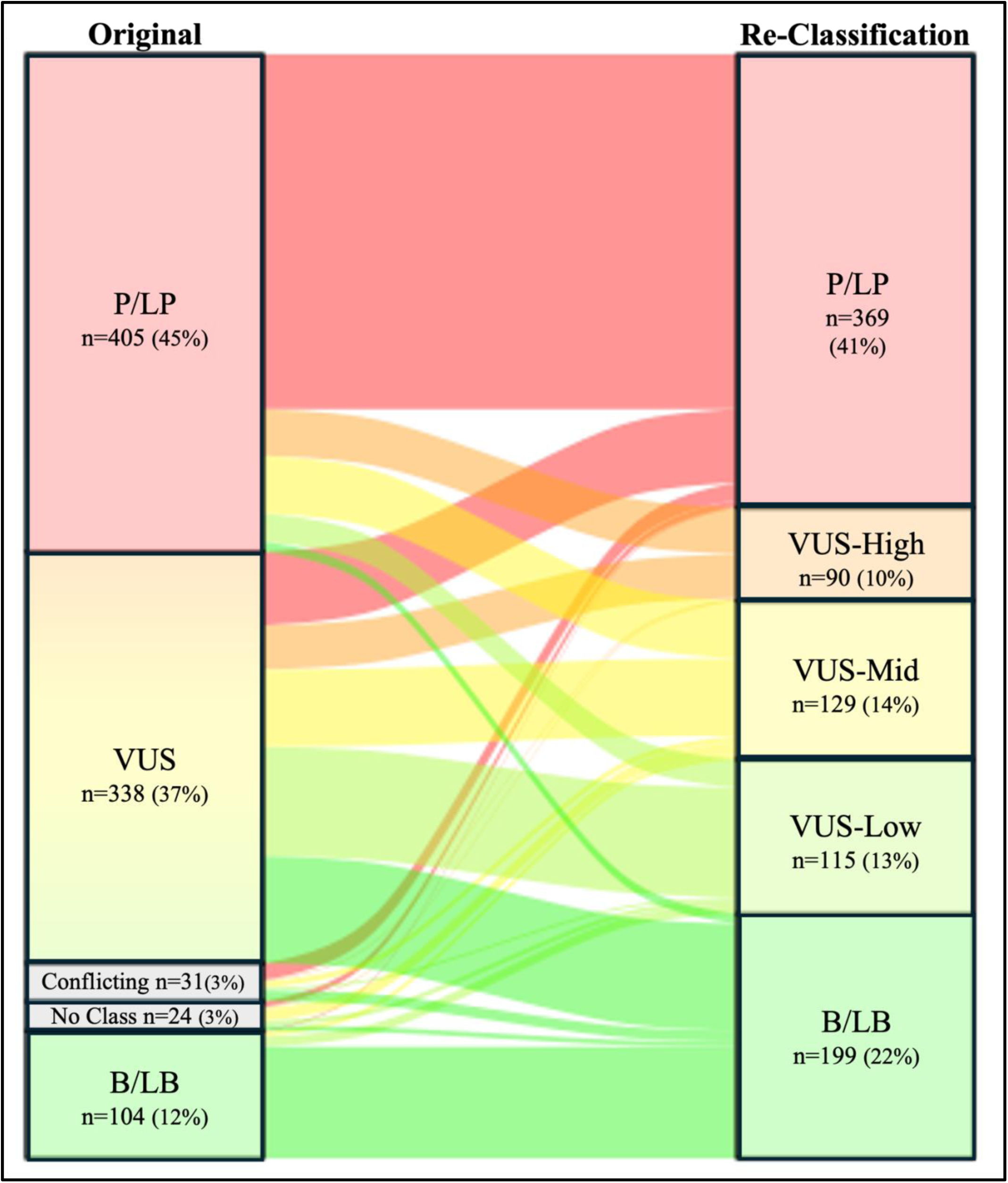
Impact of reclassification of manually curated variants. Abbreviations: P/LP, Pathogenic/Likely Pathogenic; VUS, Variant of Uncertain Significance; B/LB, Benign/Likely Benign.

### Upgrades

There were 73 upgraded variants, including 61 (18% of all reclassifications) VUS to P/LP reclassifications impacting 199 individuals clinically where this result can now guide family screening, and 12 B/LB to VUS reclassifications. Evidence driving upgrades to P/LP was mostly due to higher weighting of the ACMG/AMP criteria; PS4 proband counts, PP1 segregation with disease, and PP3 in silico tools. For example, *MYL3*:c.466G>T:p.Val156Leu, originally classified as VUS, was reclassified as P/LP by criteria PM2_sup, PS4_mod, PP3_mod(+3), equaling 6 Bayesian points. This variant is rare in healthy populations, with a total allele frequency of 0.00001363 in gnomAD v4, 9 probands reported across SHaRe, ClinVar, and the literature, and a REVEL score of 0.936 where a score ≥ 0.70 is predicting a damaging effect. The variant *MYH7*:c.5326A>G:p.Ser1776Gly was upgraded from B/LB to VUS; this variant has been seen in healthy populations, however it has increased burden in SHaRe cases compared to gnomAD controls (odds ratio [OR] 7.2; 95% CI 2.8 to 15.9) and a REVEL score of 0.776 suggesting the possibility of an intermediate effect. This highlights how evolution of variant classification criteria can better arbitrate variant status, with case-control data now the preferred method of weighting case-level evidence, though heavily reliant on availability of large patient datasets.

### Downgrades

Among the 203 downgraded variants, 108 (32% of all reclassifications) were downgraded from P/LP to VUS, impacting 196 individuals. A further 95 (29% of all reclassifications) variants were downgraded to B/LB (from P/LP or VUS). Most downgrades occurred due to new population reference datasets showing a higher than accepted allele frequency (e.g., gnomAD allele frequency precluding use of PM2_sup or BS1 being applied). Those downgraded from P/LP to B/LB (n=8) were all classified prior to the release of gnomAD v3 in 2019 and as far back as 1999 (Figure 5). Among the 237 variants classified as P/LP before 2020 that were manually curated, 73 (31%) were downgraded (Figure 6). P/LP variants downgraded to VUS were typically seen at a higher than accepted frequency in gnomAD (precluding PM2_sup use) but not at the threshold to apply benign criteria (BS1), suggesting the possibility of an intermediate effect. Alternatively, P/LP variants downgraded to VUS-High that retained rarity over time, were originally classified over a wider time interval and were typically sarcomere variants with insufficient evidence per the current ACMG criteria to reach P/LP. This reflects greater stringency in use of the criteria over time but could also point to classification by laboratories with internal clinical data that is not in the public domain.

### Reclassification of variants in sarcomere genes

Eight of the nine sarcomere genes (*TPM1, TNNT2, TNNI3, TNNC1, MYL2, MYL3, MYH7, ACTC1*) had more than 20% of variants with a reclassification (Table S1). There were 64 (10.9%) *MYBPC3* variants that changed classification. Fourteen of 203 (6.9%) variants downgraded were putative loss-of-function (high impact) variants in genes where loss-of-function is not a known mechanism of disease (Figure 3). There were 55 variants in *MYH7* downgraded, including 44/203 (21.6%) missense variants that were rare in gnomAD but lacked sufficient evidence to reach P/LP, including 22 (50%) within the PM1 (head/neck) region hot spot of *MYH7* (i.e., met PM1 but without sufficient additional evidence to reach P/LP). Conversely, 23/29 (79%) *MYH7* variants that were upgraded were within the HCM associated regional hot spot meaning PM1 could be applied as per the ClinGen CMP-EP specifications to the ACMG/AMP Variant Interpretation Guidelines for *MYH7* Version 2.0.0 (Figure 4).

**Figure 3.**
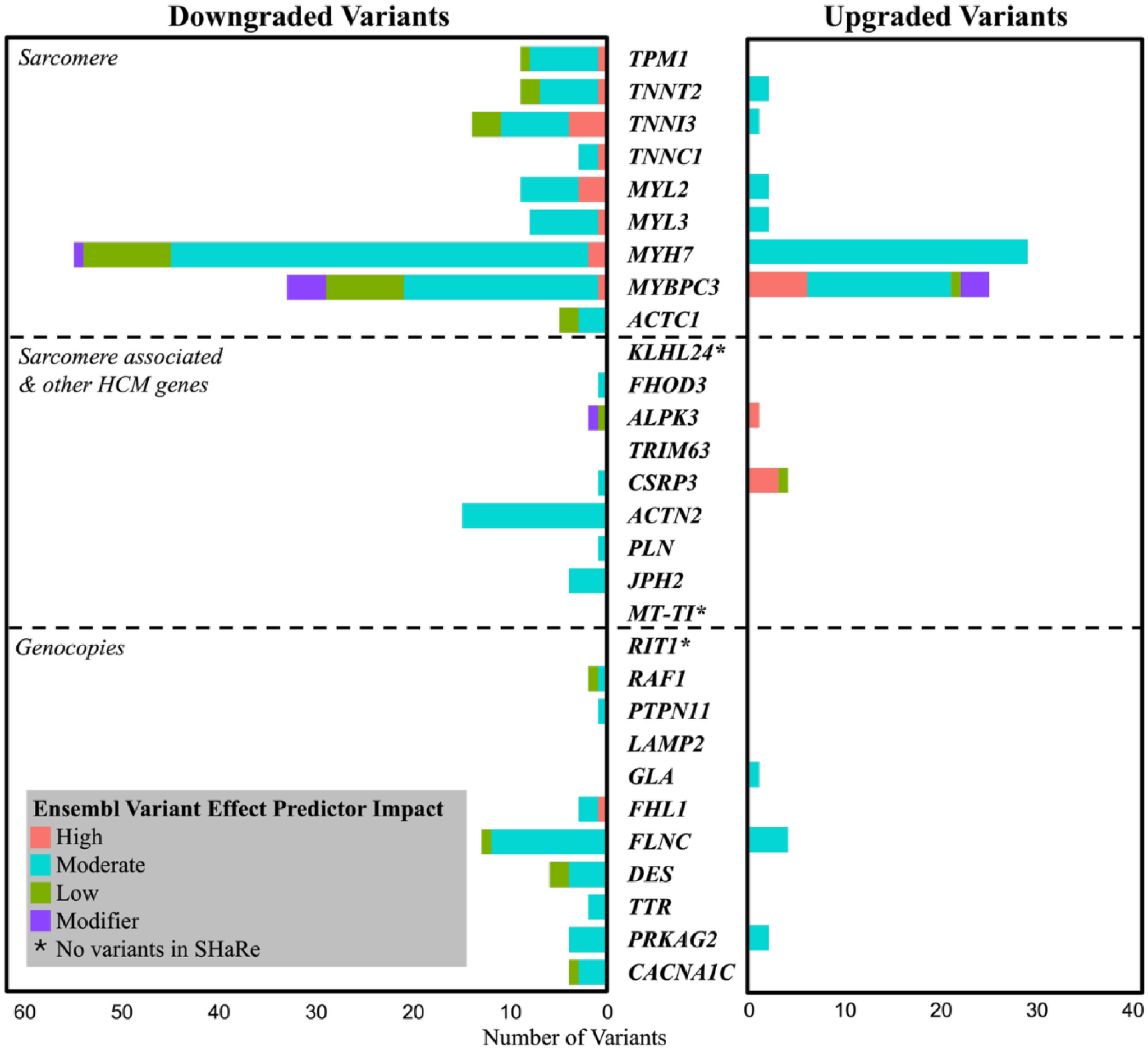
Count and predicted variant impact of variants with a change of classification for the 29 hypertrophic cardiomyopathy (HCM) genes. Predicted variant impact determined in re-annotation according to variant type by Ensembl variant effect predictor (VEP) [High (Red), Moderate (Blue), Low (green), Modifier (Purple)]. Variants that had a downgrade of classification (Left); variants that had an upgrade in classification (Right).

**Figure 4.**
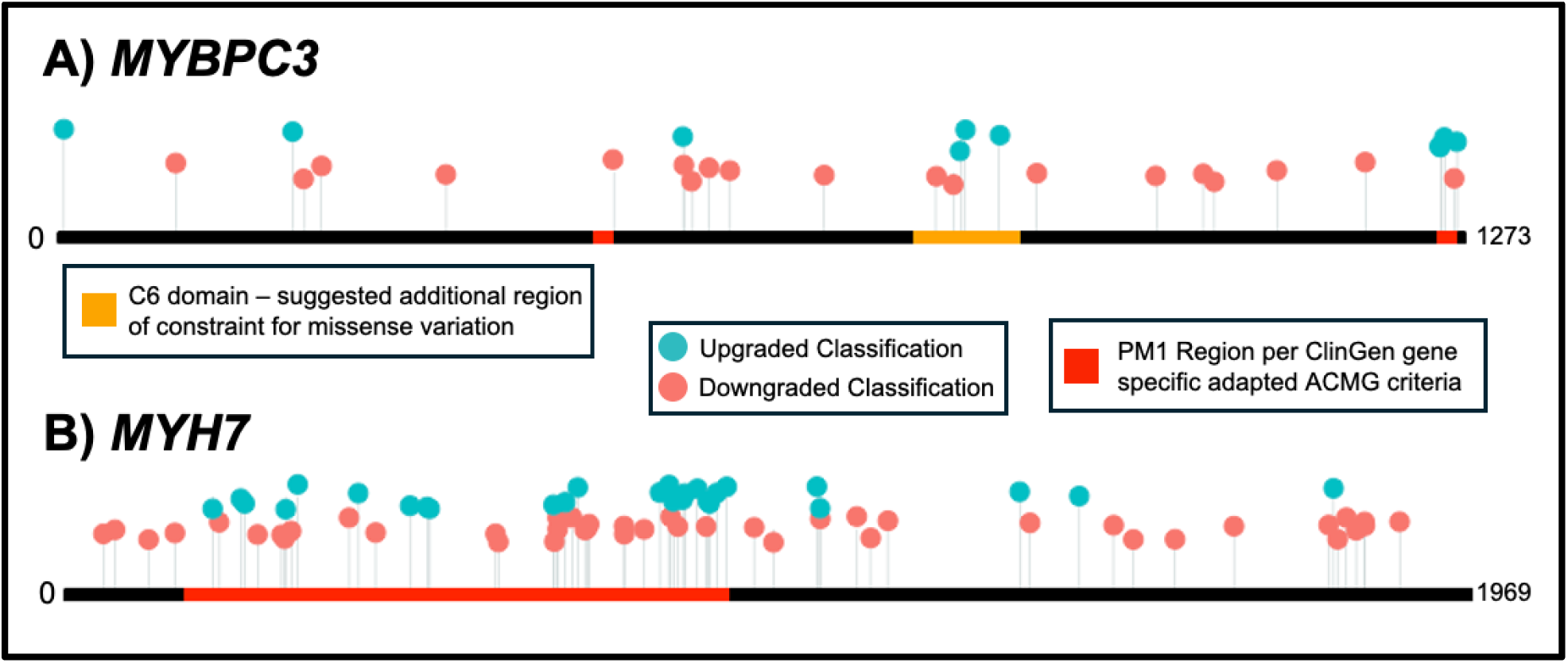
Gene topology of *MYBPC3* and *MYH7* missense variants with change of classification upon re-curation. Colored dots represent missense variants with classification changes plotted by protein position [Upgraded (Blue) and Downgraded (Red)]. **A)** MYBPC3 defined in Esemble transcript: ENSP00000442795.1 aligned by codon position starting at 0 and ending at 1273. Red PM1 regions as per ClinGen Cardiomyopathy Expert Panel Specifications to the ACMG/AMP Variant Interpretation Guidelines for MYBPC3 Version 1.0.0; codons 485-502 and 1248-1266, in orange C6 domain on MYBPC3 an additional region suggested constraint for missense variation by Helms, et al., 2020. **B)** MYH7 defined in Ensmble transcript: ENSP00000347507.3 aligned by codon position starting at 0 and ending at 1969. Red PM1 regions as per ClinGen Cardiomyopathy Expert Panel Specifications to the ACMG/AMP Variant Interpretation Guidelines for MYH7 Version 2.0.0; codons 167-931.

### Variants in genes without HCM disease association

There were an additional 1031 unique variants in genes lacking HCM disease association; 195 in genes with a limited genotype-phenotype association with HCM, 104 in syndromic genes with LVH only in the presence of overt extra-cardiac features, 676 in genes not currently associated with HCM, and 56 intergenic variants across an additional 158 genes. All variants were classified as VUS in the context of their causal role in HCM. At baseline, 5 variants in limited genes, 6 variants in syndromic genes, 73 variants in non-HCM genes, and 13 intergenic variants were considered P/LP and therefore downgraded to VUS for HCM in this review. Although the variants classified as P/LP were incidental findings in cancer and syndromic genes, therefore still P/LP for those phenotypes but not causative of HCM.

### Better characterizing uncertain variants

Sub-classification of manually curated variants resulted in fewer variants being considered a likely cause of HCM, with 90 VUS-High and 129 VUS-Mid classifications, compared to 338 (37% of all original classifications) VUS. Furthermore, following reclassification, there were 59 fewer variants considered uncertain (804 [50%] versus 863 [54%]; Table S3). The total number of B/LB variants increased from 159 (10%) at baseline to 254 (16%) after reclassification, and there were 115 VUS-Low classifications, with recent guidance recommending these either not be reported by clinical laboratories or their importance de-emphasized to patients. Taken together, 369 (40.6%) variants in total were considered either not, or probably not, HCM associated, highlighting a marked reduction in uncertainty needing to be explained to clinicians and families.

There were subtly fewer P/LP variants, with 584 (36%) at baseline and 548 (34%) upon reclassification. The number of VUS decreased in all HCM genetic subtype groups following reclassification; sarcomere variants (576 to 573), other monogenic HCM (111 to 90), and syndromic with isolated LVH/genocopy (176 to 141) (Table S3). Among sarcomere variants only, there were fewer P/LP (93% to 91%) and B/LB (81% to 67%) after reclassification, and an increase in VUS (69% to 71%).

## DISCUSSION

Genetic evidence and classification frameworks evolve quickly due to dedicated research efforts and greater availability of genomic datasets. The potential impact of this on variant reclassification is not well understood, with arbitrary time-frames provided for re-curation in disease guidelines.^2,3^ We report a real world experience of large-scale reclassification of genetic variants associated with HCM from SHaRe, a large multi-center international HCM registry. Variants were reported by clinical laboratories between 1990-2024, including 1606 variants in 29 HCM genes and a further 1031 variants in genes without HCM association. We show ∼1 in 6 (276, 17% variants) variants changed classification, and ∼1 in 10 (177, 11% variants; 395 HCM patients) were clinically important reclassifications. Sub-classification of VUS resulted in less uncertainty, with 369 (40.6%) variants reclassified as VUS-Low or B/LB, indicating a very strong probability of not being HCM associated. New evidence, specifically larger and more ancestrally diverse genomic reference databases, greater sharing of case-level and co-segregation data, and more informative *in silico* tools; or a change in frameworks, including gene-specific variant classification criteria and gene curation efforts, contributed to these changes.

Other studies have evaluated the rate of reclassification and impact on families with inherited cardiomyopathies.^17,19,25^ Numerous factors have contributed to HCM variant reclassifications. Firstly, the development of gene-specific variant classification criteria, in addition to efforts largely led by ClinGen to improve our frameworks that inform variant and gene curation, has played a key role in the reclassification of genetic variants.^23,24^ Interestingly, the defined regions of constraint for missense variations in *MYH7* and *MYBPC3,* reflected in the use of the PM1 criteria, helped to inform reclassifications but still lack specificity (Figure 4), as P/LP variants are still seen outside of hotspot regions and conversely VUS and B/LB variants are still seen within. Secondly the evolution of predictive *in silico* tools, such as REVEL,^26^ can now be used at increased weightings and may ultimately have greater benefit in ascertaining which variants are important. Lastly, the implementation of very large population reference databases and updated criteria, including a stand-alone benign criterion, contributed to B/LB variants becoming the largest group (baseline 159, 10% compared to reclassification 254, 16%). B/LB variants are not typically reported by clinical laboratories but are a common outcome of variant reclassification. Indeed, gnomAD v4, released in 2023 now comprises 730,947 exomes and 76,215 genomes from healthy individuals, including those from diverse ancestries, contributing to greater recognition of rare and common genetic variation.^21^

Updates to gene curation and better understanding mechanisms of disease have further allowed greater clarity around HCM variant reporting. The recent ClinGen HCM re-appraisal identified 5 new genes and/or inheritance patterns and changed the genotype-phenotype association of 17 genes, including three genes now considered definitive.^4,27^ Our patient population reflects real-world experience with HCM genetic testing, including many with older tests and classifications. Indeed, the speed of change in knowledge and approaches to gene and variant curation means that many genes were never previously sequenced, i.e., *ALPK3* and *FHOD3,* and that some variants are more prone to reclassification. *MYBPC3* was the only sarcomere gene with fewer than 20% of variants reclassified (at 11.1%). The development of the ClinGen CMP-EP gene specific adaptations of the ACMG/AMP criteria^24^ state that *MYBPC3* is the only gene where loss-of-function is an established mechanism of disease; as such the strongest weighted criterion, PVS1, can only be used for loss-of-function *MYBPC3* variants. Therefore, loss-of-function variants in *MYBPC3* are very unlikely to be subject to reclassification.

Recognizing the wide variability in evidence for causation among VUS, recent work has demonstrated the value of sub-classifying VUS to better reflect the likelihood of causation.^15^ Uncertain variants with the least amount of evidence, i.e., VUS-Low, have been shown to *never* be reclassified to P/LP. In fact, they were most likely to be reclassified as B/LB. Conversely, the most suspicious class of VUS, were much more likely to be classified as P/LP. We found that the VUS-High category had the fewest number of variants overall. However, identifying this subgroup of VUS can guide resource allocation, including prioritization of co-segregation studies, further clinical investigation, and functional and/or multi-omic studies to increase the body of evidence to reach P/LP. Likewise, recent advice regarding VUS sub-classification suggests VUS-low variants not to be reported or be de-emphasized to patients knowing that they are extremely unlikely to be causal. Approximately half of the VUS that underwent manual curation were eventually considered VUS-Low or B/LB, highlighting the marked time and resource reduction that could be possible using this approach.

Sub-classification of VUS can allow for greater clinical impact when reclassifying variants. For example, the variant *MYH7* c.350A>T p.Tyr117Phe originally classified as P/LP in 2017 and downgraded to VUS-Low upon re-classification, means the variant can indeed be de-emphasized as the causative finding to both the clinician and the patient. On the other hand, the variant *MYH7* c.3170G>A p.Gly1057Asp originally classified as P/LP in 2017 and downgraded to VUS-High following reclassification, highlights this is still likely to be a causative variant but currently there is a lack of supportive evidence. Efforts to convey this nuance to the family and consider how additional supportive evidence may be gleaned would be extremely worthwhile in this situation.

A key finding from this study was the importance of proband information. We show an example of four VUS-High that underwent reclassification and with the inclusion of internal SHaRe proband data, these variants could be upgraded to P/LP, impacting 13 patients in SHaRe. While clinical laboratories maintain comprehensive internal variant databases, we highlight that large-scale public data sharing is critical for providing evidence for some variant classifications. Indeed, the benefit of data sharing in improving classification accuracy has been reported previously.^28,29^ Public repositories such as ClinVar have transformed our ability to share case-level data, with many clinical laboratories now submitting their variant classifications, though ongoing challenges include lack of phenotype data, ancestry of cases and inability to discern duplicate case reports (e.g., a patient reported by multiple laboratories, or overlap between a literature and laboratory report). In addition, for variants in genes such as *MYH7*, recognition of whether the available evidence supports an HCM or a DCM phenotype remains important, therefore case reporting without phenotype data is not necessarily helpful. Of course, there are confidentiality issues with public sharing of variant evidence including case-level details, however these must be explored to ensure we continue to minimize ongoing uncertainty, and at worst, variant misclassification.

Recognizing this need, we have made these HCM variant classifications publicly accessible in a custom-developed variant browser (https://www.cardiodb.org/share_browser/). This includes, for each variant where available; GRCh38 location, variant nomenclature (canonical transcript), whether within established hotspot of gene, proband count, family member count, zygosity/allele count, ancestry, case burden (odds ratio & 95% confidence interval of cases within SHaRe versus gnomAD), gnomAD frequencies (including sub-populations) and allele counts, in silico tool scores (REVEL, spliceAI), and SHaRe classification.

Reclassification of genetic variants remains an ongoing challenge. While guidelines recommend this be performed, there is little evidence to guide who takes responsibility or how often reevaluation should occur.^2^ At present, genetic testing is treated as a one-time test. However, with the pace of new knowledge, this paradigm should be reconsidered. For many patients, genetic results are simply not static and there is need for periodic reclassification and/or re-testing over time. Against a backdrop of substantial global barriers to accessing genomic services, demand is expected to increase further as targeted therapies become more widely available. For those who undertake their regular follow-up in centers of expertise, including with access to experienced cardiac genetic counselors, this is likely to pose an ideal timepoint for periodic reclassification.^8,30^ However, the large majority of HCM patients globally do not attend specialized centers. Many clinical laboratories now have established pathways to automatically notify clinicians of a variant reclassification, posing a significant burden on busy and often non-expert clinicians. Fortunately, as we have shown, better knowledge and improved frameworks have resulted in less uncertainty, therefore the impact of further reclassifications over time should be less (Figure 5).

**Figure 5.**
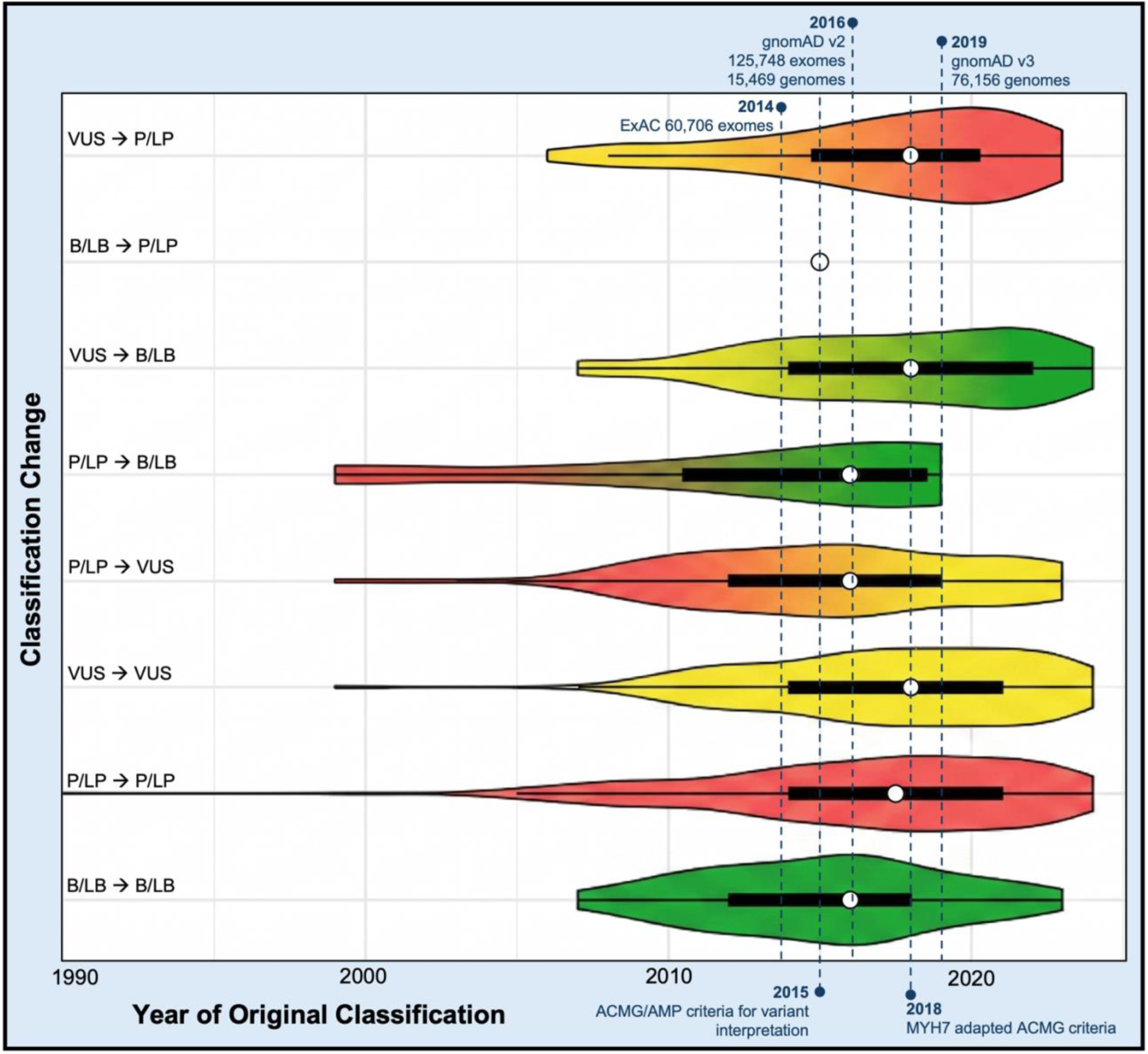
Temporal distribution of classification changes relative to major genomic milestones. Each violin illustrates the density and temporal distribution of variants for a given classification transition. The plots feature a color gradient corresponding to the clinical severity of the classification (red for P/LP, yellow for VUS, green for B/LB). Embedded within each violin is a standard box plot where the white dot indicates the median year of original classification, and the thick black bar represents the interquartile range (IQR).

Building on these observations, we propose a simple workflow to guide reclassification with a goal to reduce uncertainty (Figure 6). The temporal cut-offs applied in this workflow reflect major expansions of population reference data, most notably the release of gnomAD v3 in 2019, which substantially increased the resolution of allele frequency estimates across ancestries and precipitated many of the downgrades. P/LP variants classified prior to 2020 warrant re-curation as a priority, with 31% of manually curated variants (i.e., variants with new evidence) were downgraded in our cohort, most commonly due to updated population allele frequency data. Prioritization of VUS re-curation should be guided by the pre-test probability of a heritable cause, including family history of HCM and clinical features suggestive of a monogenic etiology. Concerted efforts to understand the genetic architecture of HCM will continue to mean previous HCM genetic testing may need to be revisited. While we work to translate our research discoveries to clinical practice, reconciling what this means for previously tested patients should be front of mind. Development of approaches aimed at educating and supporting clinicians to know when reclassification or re-testing is appropriate is critically important and should form part of the accepted HCM patient management pathways. Further, publicly available educational materials to guide patients themselves could empower individuals to seek reevaluation of genetic test results where appropriate.

**Figure 6.**
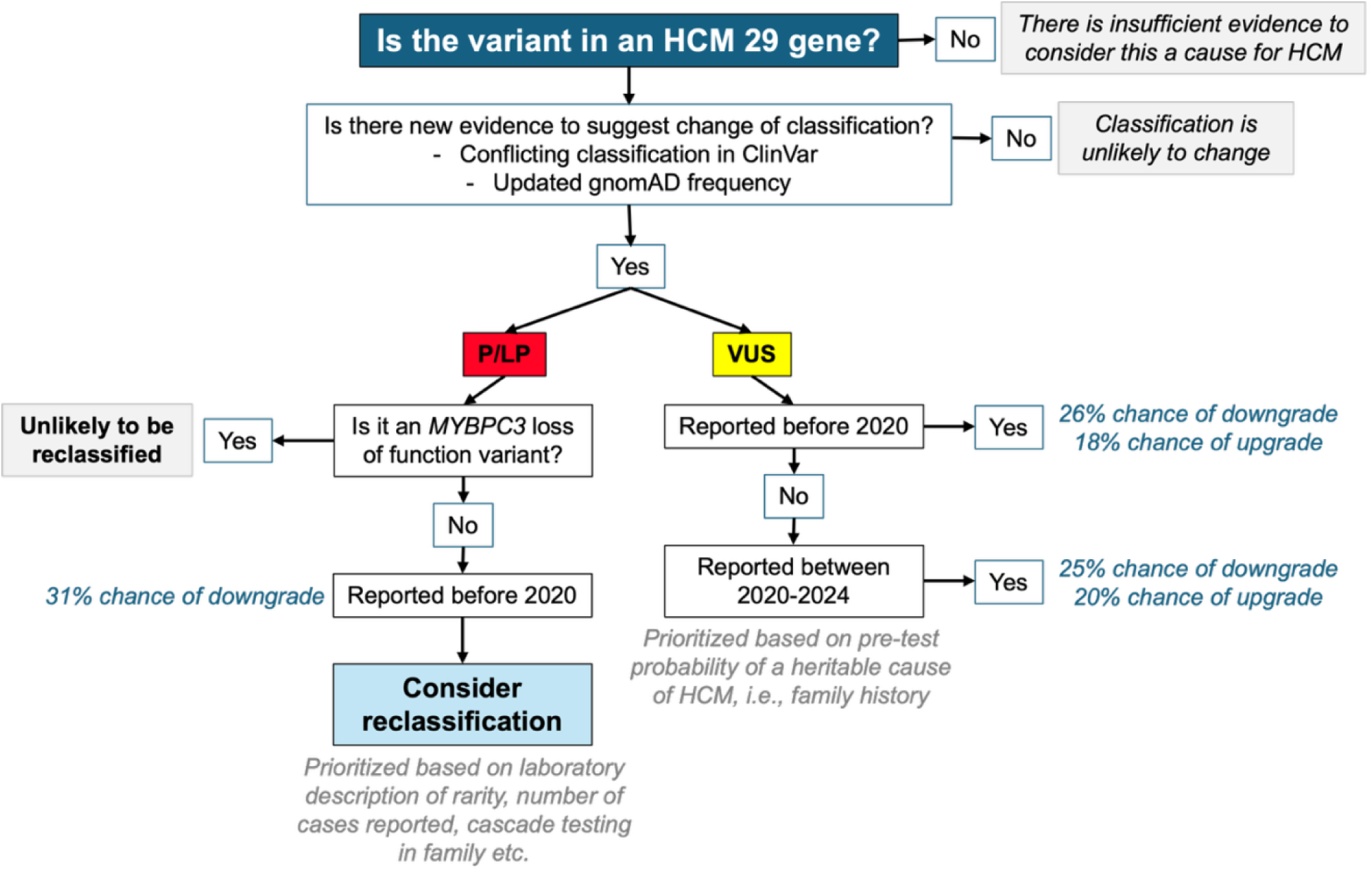
HCM variant reclassification workflow. Proposed HCM variant reclassification framework to guide clinical and research prioritization of variant reclassification. Percentage chance of classification change calculated from variants in the manual curation group after computational re-annotation.

## CONCLUSION

Classification of genetic variants reflect our knowledge and practices at a point in time. Using current classification algorithms and all available clinical data, one in ten HCM variants in SHaRe had a clinically meaningful reclassification. Further, sub-classification of VUS reduced uncertainty, enabling streamlined genetic counseling discussions. As we continue to push forward on HCM genetic discovery and translation to clinical practice, efforts to inform what this means for previously tested patients must be considered.

## Data Availability

All data produced in the present study are available upon reasonable request to the authors

https://www.cardiodb.org/share_browser/#/

## Nonstandard Abbreviations and Acronyms

HCM: Hypertrophic Cardiomyopathy
SHaRe: Sarcomeric Human Cardiomyopathy Registry
LVH Left: Ventricular Hypertrophy
ACMG: American College of Medical Genetics and Genomics
P/LP: Pathogenic/Likely Pathogenic
VUS: Variant of Uncertain Significance
B/LB: Benign/Likely Benign
VEP: Variant Effect Predictor
LOF: Loss-of-function
AD: Autosomal dominant
ClinGen CMP-EP: Clinical Genome Resource Cardiomyopathy Expert Panel

## Ethics statement

Each participating site has received ethics approval in accordance with local policies, as per the following: ethical approval requiring informed consent was obtained from Cincinnati Children’s Hospital USA; Children’s Hospital of Philadelphia, USA; Michigan Medical, USA; Yale Medical, USA; Royal Brompton Hospital, United Kingdom; Erasmus University Medical Center, The Netherlands; Florence Centre for Cardiomyopathies, Italy; Sydney Local Health District Royal Prince Alfred Hospital Australia; and InCor, Heart Institute, University of Sao Paulo, Brazil ethics committees. Waiver of consent was granted by Stanford School of Medicine, USA; Brigham and Women’s Hospital, USA; Boston Children’s Hospital, USA; Pennsylvania University Medical Center USA, and Sydney Local Health District Royal Prince Alfred Hospital Australia ethics committees.

## SUPPLEMENTARY MATERIAL

**Table S1.**
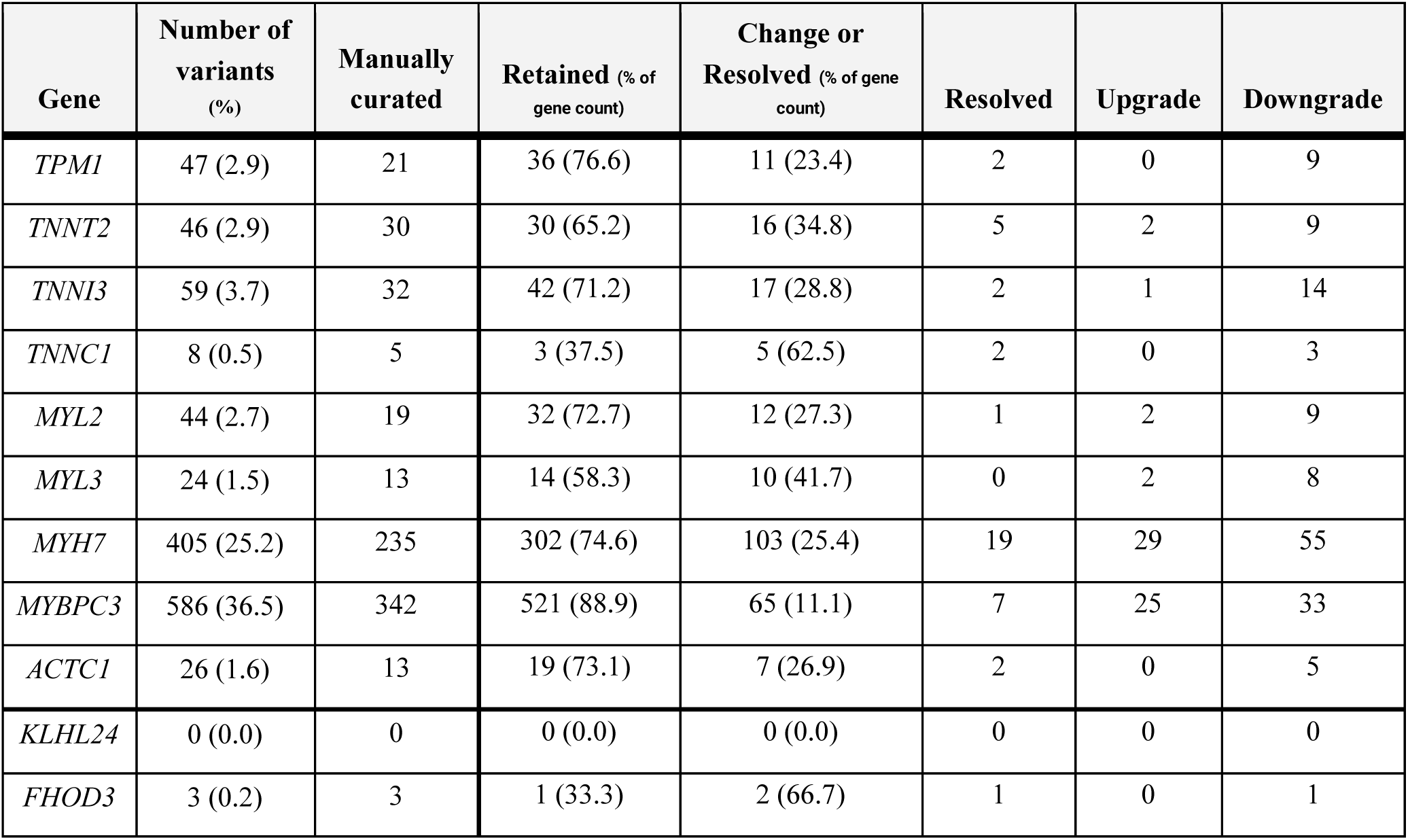
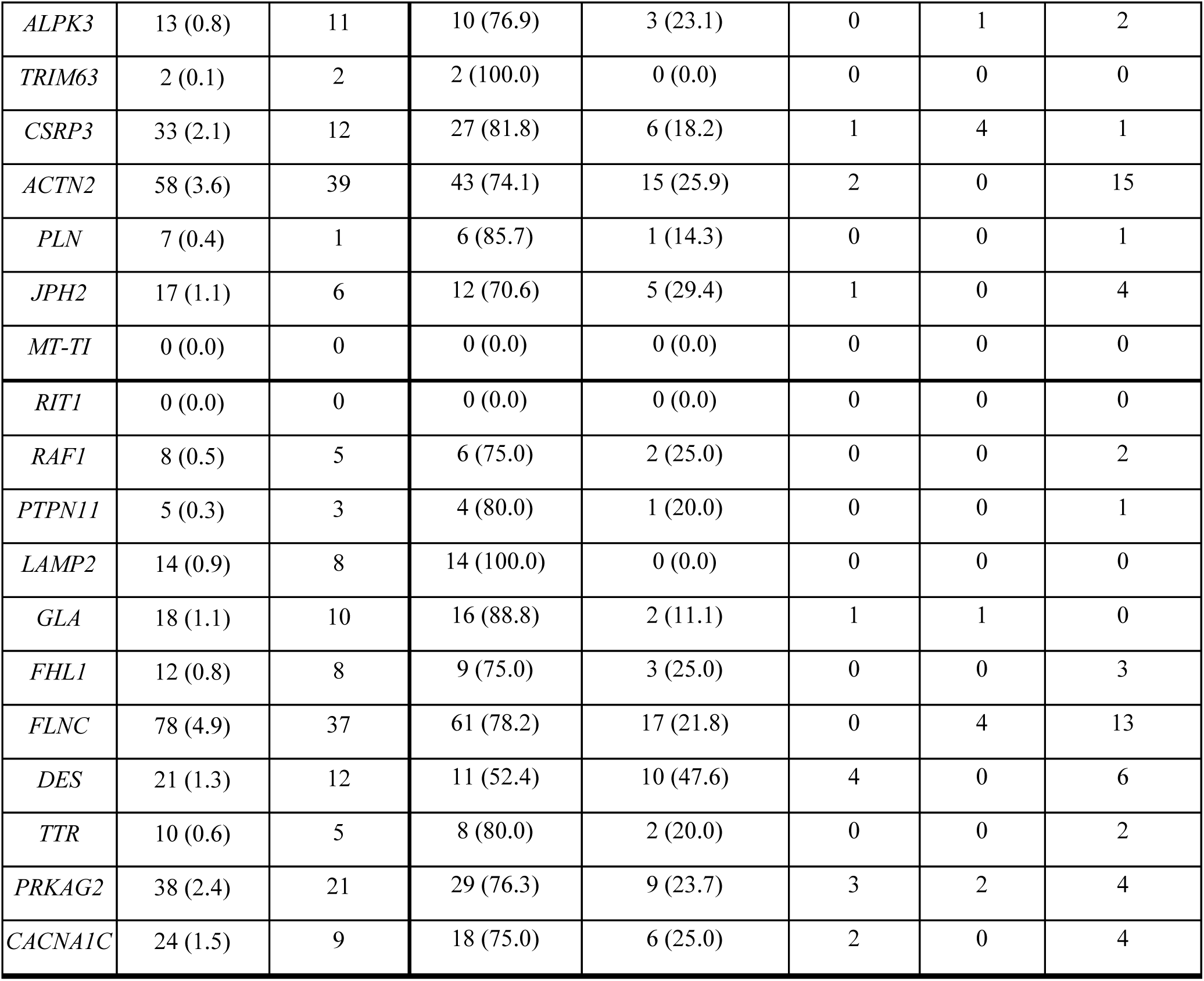
Reclassification of genetic variants in HCM genes.

| Gene | Number of variants (%) | Manually curated | Retained (% of gene count) | Change or Resolved (% of gene count) | Resolved | Upgrade | Downgrade |
| --- | --- | --- | --- | --- | --- | --- | --- |
| TPM1 | 47 (2.9) | 21 | 36 (76.6) | 11 (23.4) | 2 | 0 | 9 |
| TNNT2 | 46 (2.9) | 30 | 30 (65.2) | 16 (34.8) | 5 | 2 | 9 |
| TNNI3 | 59 (3.7) | 32 | 42 (71.2) | 17 (28.8) | 2 | 1 | 14 |
| TNNC1 | 8 (0.5) | 5 | 3 (37.5) | 5 (62.5) | 2 | 0 | 3 |
| MYL2 | 44 (2.7) | 19 | 32 (72.7) | 12 (27.3) | 1 | 2 | 9 |
| MYL3 | 24 (1.5) | 13 | 14 (58.3) | 10 (41.7) | 0 | 2 | 8 |
| MYH7 | 405 (25.2) | 235 | 302 (74.6) | 103 (25.4) | 19 | 29 | 55 |
| MYBPC3 | 586 (36.5) | 342 | 521 (88.9) | 65 (11.1) | 7 | 25 | 33 |
| ACTC1 | 26 (1.6) | 13 | 19 (73.1) | 7 (26.9) | 2 | 0 | 5 |
| KLHL24 | 0 (0.0) | 0 | 0 (0.0) | 0 (0.0) | 0 | 0 | 0 |
| FHOD3 | 3 (0.2) | 3 | 1 (33.3) | 2 (66.7) | 1 | 0 | 1 |
| *ALPK3* | 13 (0.8) | 11 | 10 (76.9) | 3 (23.1) | 0 | 1 | 2 |
| *TRIM63* | 2 (0.1) | 2 | 2 (100.0) | 0 (0.0) | 0 | 0 | 0 |
| *CSRP3* | 33 (2.1) | 12 | 27 (81.8) | 6 (18.2) | 1 | 4 | 1 |
| *ACTN2* | 58 (3.6) | 39 | 43 (74.1) | 15 (25.9) | 2 | 0 | 15 |
| *PLN* | 7 (0.4) | 1 | 6 (85.7) | 1 (14.3) | 0 | 0 | 1 |
| *JPH2* | 17 (1.1) | 6 | 12 (70.6) | 5 (29.4) | 1 | 0 | 4 |
| *MT-TI* | 0 (0.0) | 0 | 0 (0.0) | 0 (0.0) | 0 | 0 | 0 |
| *RIT1* | 0 (0.0) | 0 | 0 (0.0) | 0 (0.0) | 0 | 0 | 0 |
| *RAF1* | 8 (0.5) | 5 | 6 (75.0) | 2 (25.0) | 0 | 0 | 2 |
| *PTPN11* | 5 (0.3) | 3 | 4 (80.0) | 1 (20.0) | 0 | 0 | 1 |
| *LAMP2* | 14 (0.9) | 8 | 14 (100.0) | 0 (0.0) | 0 | 0 | 0 |
| *GLA* | 18 (1.1) | 10 | 16 (88.8) | 2 (11.1) | 1 | 1 | 0 |
| *FHL1* | 12 (0.8) | 8 | 9 (75.0) | 3 (25.0) | 0 | 0 | 3 |
| *FLNC* | 78 (4.9) | 37 | 61 (78.2) | 17 (21.8) | 0 | 4 | 13 |
| *DES* | 21 (1.3) | 12 | 11 (52.4) | 10 (47.6) | 4 | 0 | 6 |
| *TTR* | 10 (0.6) | 5 | 8 (80.0) | 2 (20.0) | 0 | 0 | 2 |
| *PRKAG2* | 38 (2.4) | 21 | 29 (76.3) | 9 (23.7) | 3 | 2 | 4 |
| *CACNA1C* | 24 (1.5) | 9 | 18 (75.0) | 6 (25.0) | 2 | 0 | 4 |

**Table S2.** Variants resolved from conflicting classifications within SHaRe.

| Gene | HGVSc | HGVSp | SHaRe Allele Count (n) | gnomAD Allele Count (n) | Baseline Class | Final Class |
| --- | --- | --- | --- | --- | --- | --- |
| MYBPC3 | c.706A>G | p.Ser236Gly | 8 | 189977 | LB/VUS | B/LB |
| MYH7 | c.4472C>G | p.Ser1491Cys | 2 | 18642 | P/VUS | B/LB |
| MYBPC3 | c.565G>A | p.Val189Ile | 8 | 4062 | LB/VUS | B/LB |
| TNNT2 | c.862C>T | p.Arg288Cys | 21 | 827 | LB/LP/VUS | B/LB |
| GLA | c.352C>T | p.Arg118Cys | 2 | 732 | LB/VUS | B/LB |
| TNNT2 | c.692T>C | p.Ile231Thr | 2 | 249 | LB/VUS | B/LB |
| ACTN2 | c.1930G>A | p.Ala644Thr | 2 | 237 | LB/VUS | B/LB |
| MYH7 | c.115G>A | p.Val39Met | 3 | 98 | P/VUS | B/LB |
| TNNT2 | c.887G>A | p.Arg296His | 2 | 46 | LB/VUS | B/LB |
| MYH7 | c.5561C>T | p.Thr1854Met | 2 | 45 | LP/VUS | P/LP |
| TNNC1 | c.430A>G | p.Asn144Asp | 7 | 31 | LP/VUS | VUS |
| MYBPC3 | c.1224-80G>A | - | 2 | 30 | LP/VUS | P/LP |
| MYH7 | c.3134G>T | p.Arg1045Leu | 4 | 24 | LP/VUS | P/LP |
| TNNC1 | c.23C>T | p.Ala8Val | 2 | 20 | LP/VUS | P/LP |
| MYH7 | c.2080C>T | p.Arg694Cys | 18 | 15 | LP/P/VUS | P/LP |
| MYH7 | c.4259G>A | p.Arg1420Gln | 3 | 10 | LP/VUS | P/LP |
| MYH7 | c.611G>T | p.Arg204Leu | 2 | 6 | P/VUS | VUS |
| TPM1 | c.82G>C | p.Asp28His | 2 | 3 | LP/VUS | VUS |
| TPM1 | c.574G>A | p.Glu192Lys | 6 | 3 | P/VUS | P/LP |
| MYH7 | c.2012G>A | p.Arg671His | 3 | 2 | LP/VUS | P/LP |
| MYBPC3 | c.481C>A | p.Pro161Thr | 2 | 2 | LP/VUS | P/LP |
| MYH7 | c.3346G>A | p.Glu1116Lys | 6 | 1 | P/VUS | VUS |
| PRKAG2 | c.1004T>C | p.Met335Thr | 4 | 1 | LP/VUS | VUS |
| TNNI3 | c.356C>A | p.Thr119Asn | 1 | 1 | LP/VUS | VUS |
| TNNT2 | c.305G>A | p.Arg102Gln | 6 | 1 | P/VUS | P/LP |
| MYH7 | c.1051A>G | p.Lys351Glu | 3 | 0 | LP/P/VUS | P/LP |
| MYH7 | c.4402G>C | p.Glu1468Gln | 5 | 0 | LP/VUS | VUS |
| MYH7 | c.2727C>G | p.Ile909Met | 4 | 0 | LP/VUS | P/LP |
| MYH7 | c.2306T>C | p.Leu769Pro | 2 | 0 | LP/VUS | P/LP |
| MYBPC3 | c.1096C>T | p.Gln366Ter | 2 | 0 | P/VUS | P/LP |
| MYH7 | c.1228T>C | p.Tyr410His | 2 | 0 | P/VUS | VUS |

**Table S3.** Overview of baseline and reclassified HCM variants.

| HCM genetic subtype | P/LP |  | VUS |  | B/LB |  |
| --- | --- | --- | --- | --- | --- | --- |
|  | Baseline (% of classification) | Final (% of classification) | Baseline (% of classification) | Final (% of classification) | Baseline (% of classification) | Final (% of classification) |
| Sarcomere | 541 (93%) | 501 (91%) | 576 (69%) | 573 (71%) | 128 (81%) | 171 (67%) |
| Sarcomere-associated & other monogenic HCM | 13 (2%) | 14 (3%) | 111 (13%) | 90 (11%) | 9 (6%) | 29 (11%) |
| Syndromic causing isolated LVH/HCM Genocopy | 30 (5%) | 33 (6%) | 176 (20%) | 141 (18%) | 22 (14%) | 54 (21%) |
| Total (% of total variants) | 584 (36%) | 548 (34%) | 863 (54%) | 804 (50%) | 159 (10%) | 254 (16%) |

